## Supplementary materials for "Crykey: Rapid Identification of SARS-CoV-2 Cryptic Mutations in Wastewater"

### Supplementary Figures

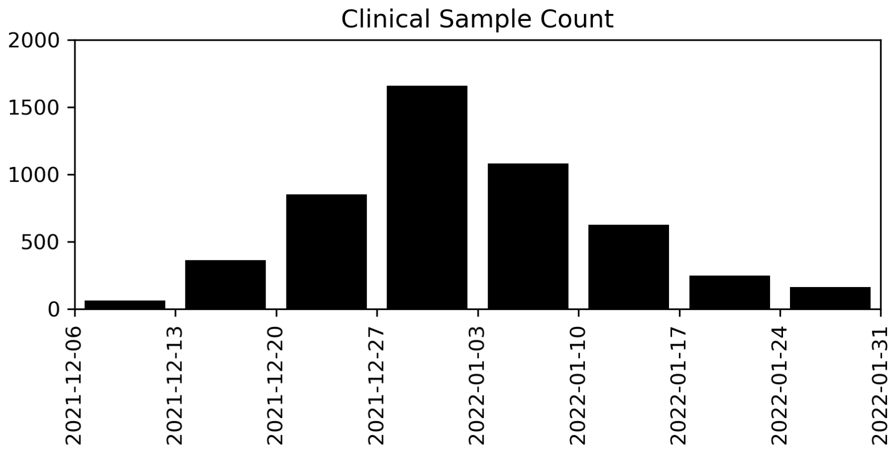


**Supplementary Figure 1. Number of clinical samples used for CR detection during the start of the Omicron season in Houston.** 5,060 sequenced clinical samples collected between 2021-12-06 and 2022-01-31. Each bar represents the number of samples collected during the one week period. The sample count shows a normal distribution pattern where the peak occurs in the week between 2021-12-27 and 2022-01-03.

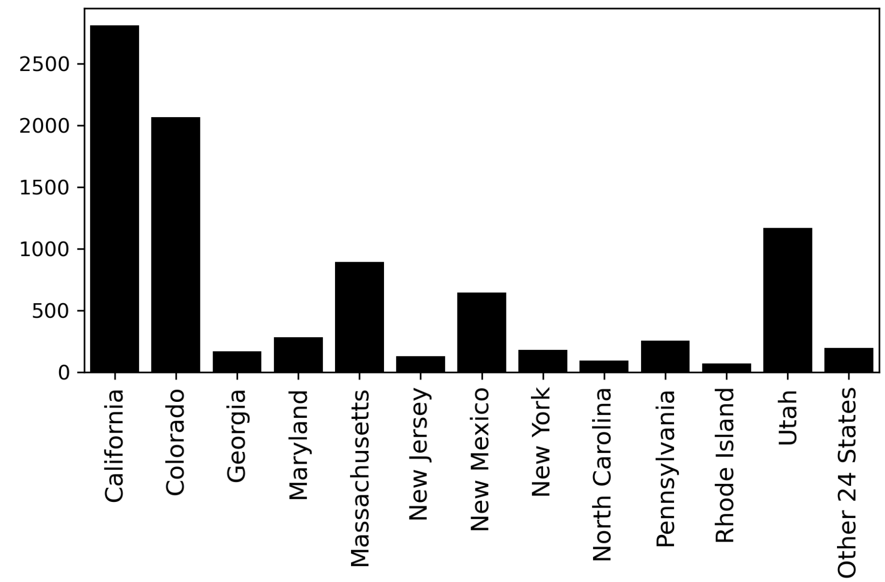


**Supplementary Figure 2. Count of clinical samples used for CR detection during the start of the Omicron season in non-Texas states from the US.** 8,969 sequenced clinical samples were collected between 2021-12-06 and 2022-01-31.

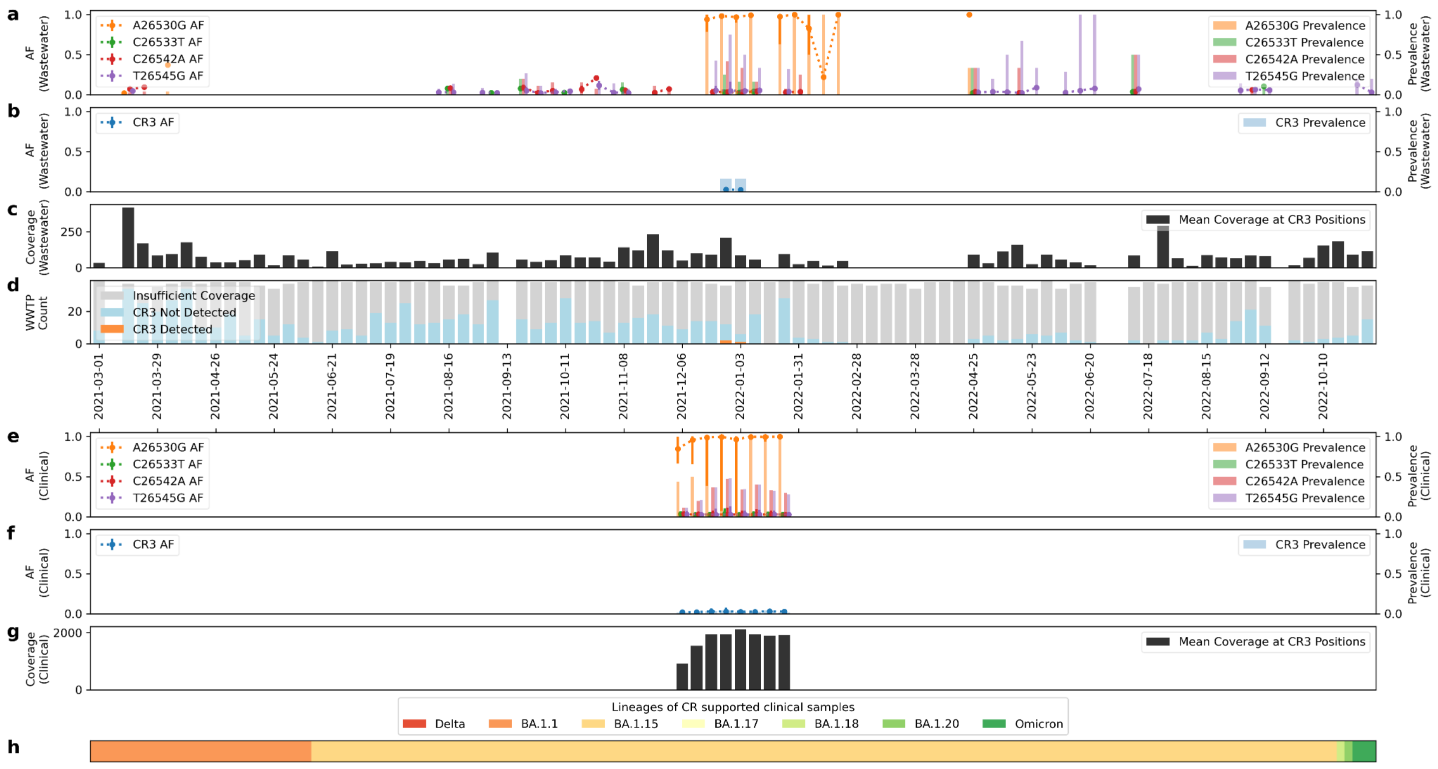


**Supplementary Figure 3. CR3 detected in wastewater and clinical samples from Greater Houston.** Figure a-d are information of CR in wastewater each week. a) shows the individual AF (with mean AF shown in dotted line, minimum/maximum of AF observed shown as error bars, the same below) and prevalence rate (shown as bars, the same below) of mutations within CR. b) shows the AF and prevalence rate of CR. c) shows the mean coverage at CR5 locations. d) shows the sample qualities and Crykey detections, with samples of insufficient coverage colored in gray, samples of CR absent colored in blue, and samples of CR detected colored in orange. Figure e-h are information of CR in clinical samples of Houston for 8 weeks of sampling period. e) shows the individual AF and prevalence rate of mutations within CR. f) shows AF and prevalence rate of CR. g) shows the mean coverage at CR locations. h) shows the distribution of the PANGO lineages of the consensus genomes of the clinical samples with CR. Delta genomes are not found in any of the samples. All Omicron genomes other than BA.1.1, BA.1.15, BA.1.17, BA.1.18, BA.1.20 are combined and denoted as Omicron. For figure a-c, and e-h, wastewater and clinical samples with insufficient coverage (<10x at CR location) are excluded from the analysis.

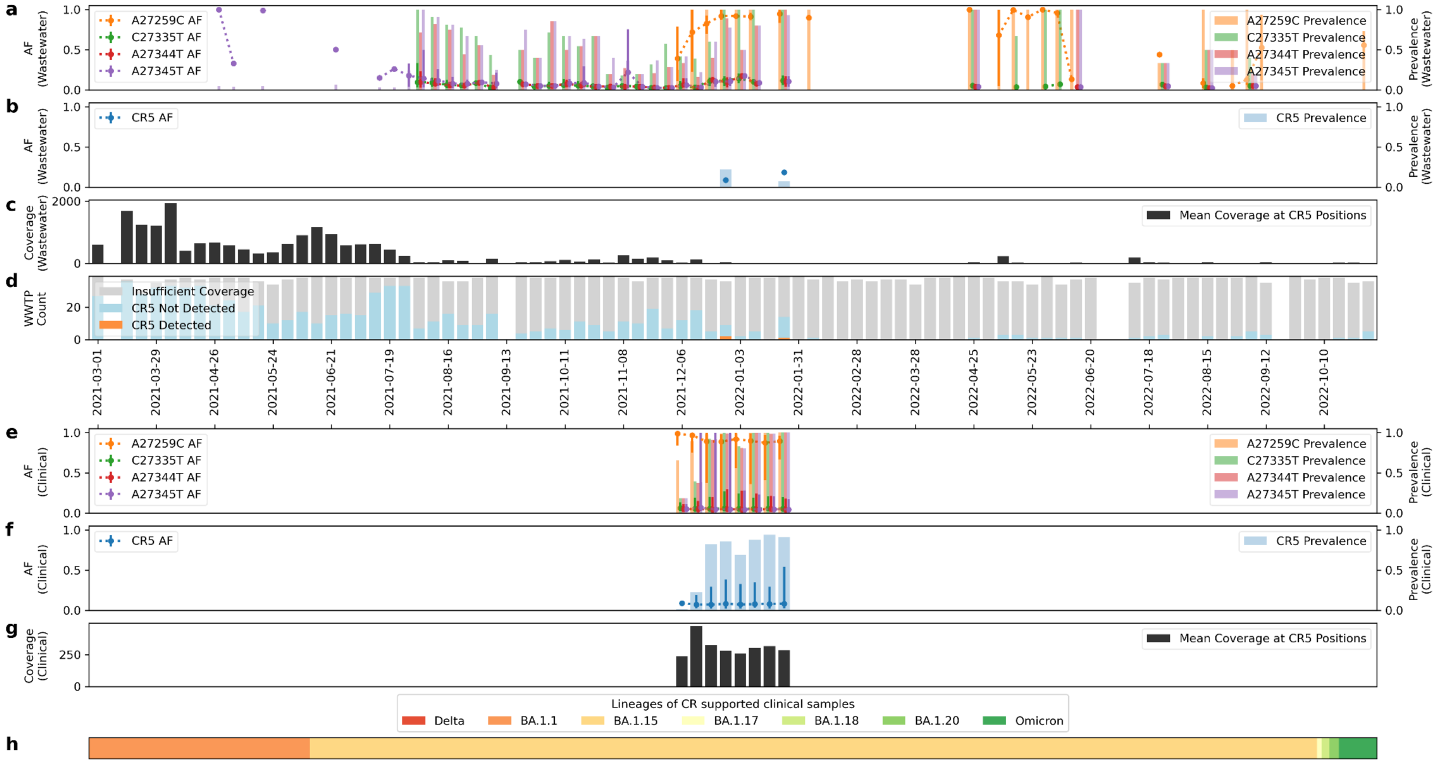


**Supplementary Figure 4. CR5 detected in wastewater and clinical samples from Greater Houston.** Figure a-d are information of CR in wastewater each week. a) shows the individual AF (with mean AF shown in dotted line, minimum/maximum of AF observed shown as error bars, the same below) and prevalence rate (shown as bars, the same below) of mutations within CR. b) shows the AF and prevalence rate of CR. c) shows the mean coverage at CR5 locations. d) shows the sample qualities and Crykey detections, with samples of insufficient coverage colored in gray, samples of CR absent colored in blue, and samples of CR detected colored in orange. Figure e-h are information of CR in clinical samples of Houston for 8 weeks of sampling period. e) shows the individual AF and prevalence rate of mutations within CR. f) shows AF and prevalence rate of CR. g) shows the mean coverage at CR locations. h) shows the distribution of the PANGO lineages of the consensus genomes of the clinical samples with CR. Delta genomes are not found in any of the samples. All Omicron genomes other than BA.1.1, BA.1.15, BA.1.17, BA.1.18, BA.1.20 are combined and denoted as Omicron. For figure a-c, and e-h, wastewater and clinical samples with insufficient coverage (<10x at CR location) are excluded from the analysis.

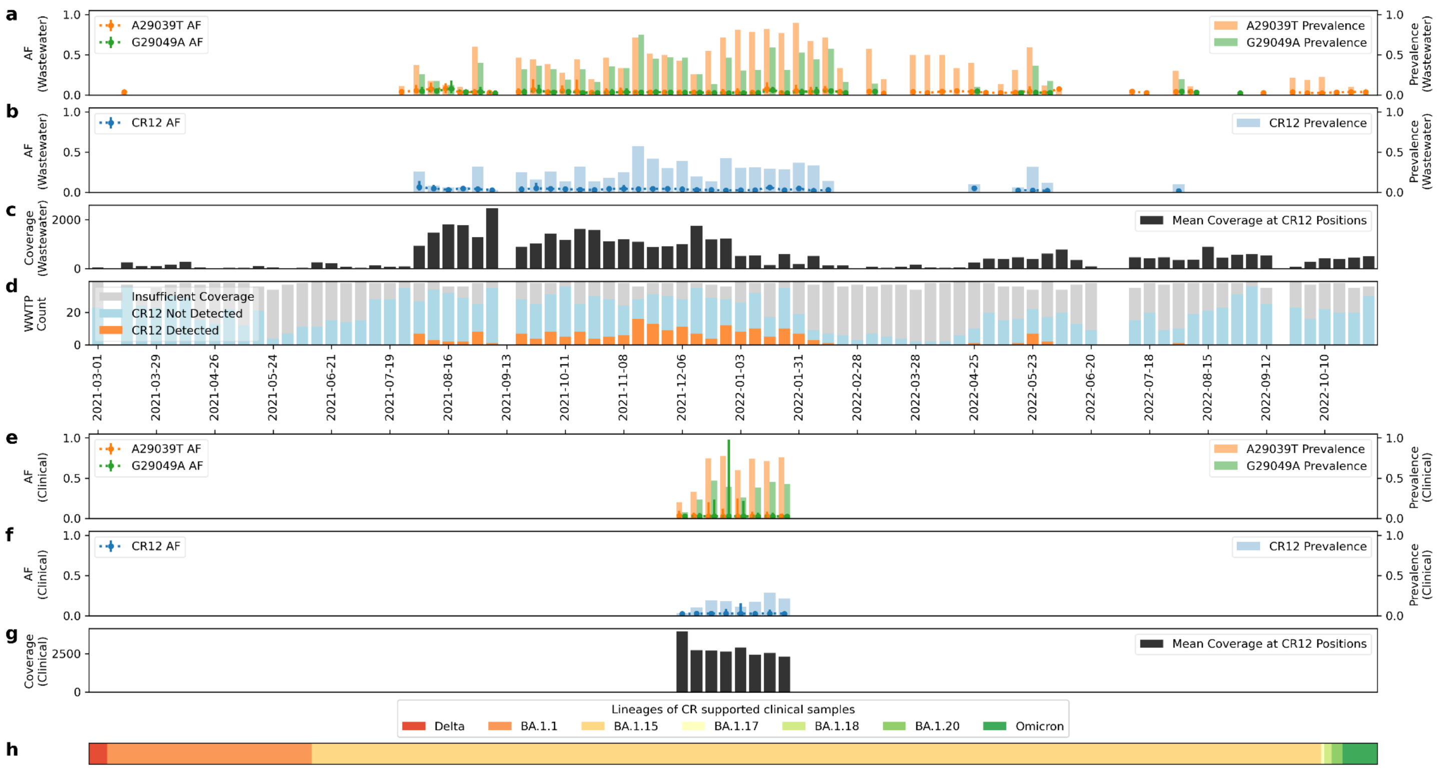


**Supplementary Figure 5. CR12 detected in wastewater and clinical samples from Greater Houston.** Figure a-d are information of CR in wastewater each week. a) shows the individual AF (with mean AF shown in dotted line, minimum/maximum of AF observed shown as error bars, the same below) and prevalence rate (shown as bars,  the same below) of mutations within CR. b) shows the AF and prevalence rate of CR. c) shows the mean coverage at CR5 locations. d) shows the sample qualities and Crykey detections, with samples of insufficient coverage colored in gray, samples of CR absent colored in blue, and samples of CR detected colored in orange. Figure e-h are information of CR in clinical samples of Houston for 8 weeks of sampling period. e) shows the individual AF and prevalence rate of mutations within CR. f) shows AF and prevalence rate of CR. g) shows the mean coverage at CR locations. h) shows the distribution of the PANGO lineages of the consensus genomes of the clinical samples with CR. Delta genomes are not found in any of the samples. All Omicron genomes other than BA.1.1, BA.1.15, BA.1.17, BA.1.18, BA.1.20 are combined and denoted as Omicron. For figure a-c, and e-h, wastewater and clinical samples with insufficient coverage (<10x at CR location) are excluded from the analysis.

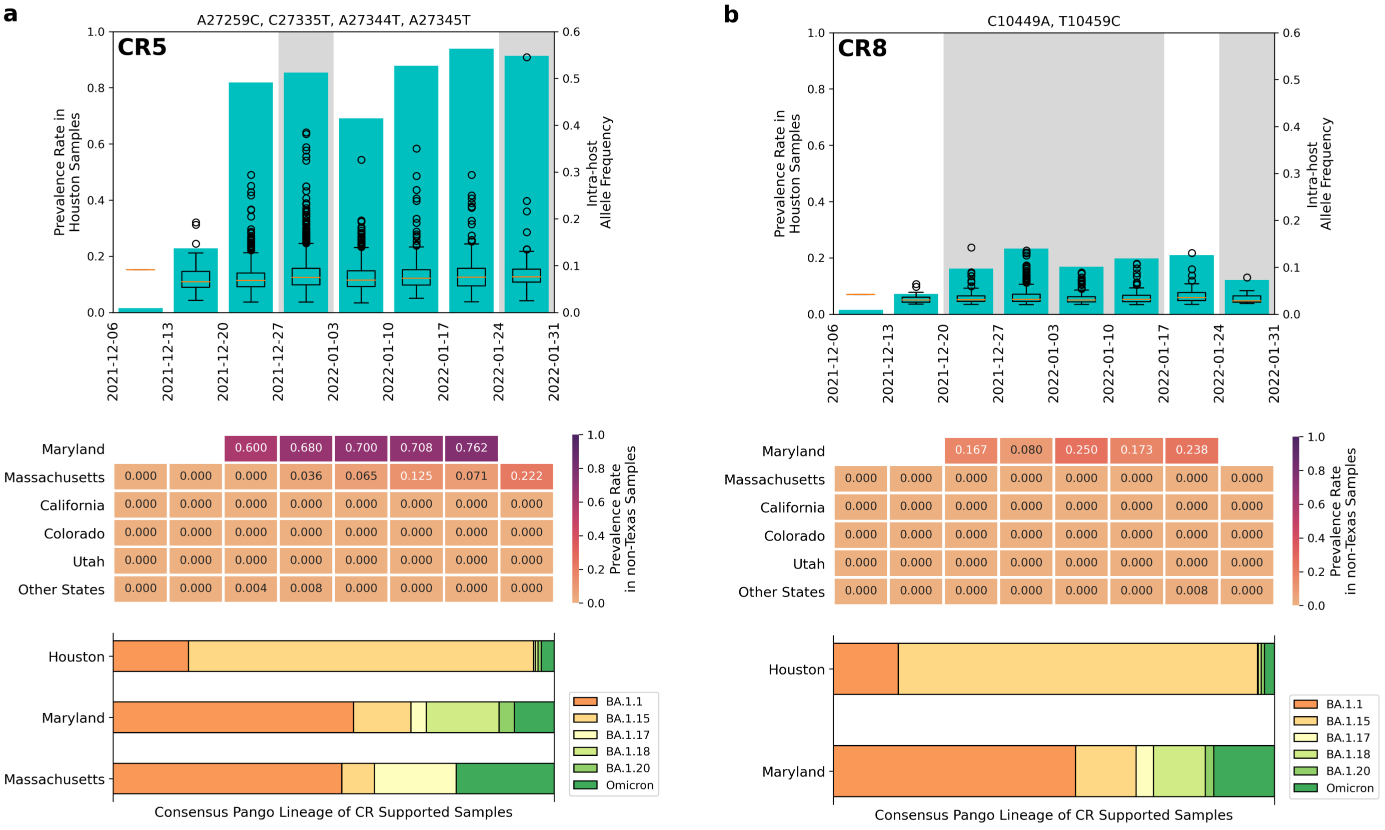


**Supplementary Figure 6. CRs detected in clinical samples from Houston and other US cities.** The figure shows detailed information of a) CR A27259C-C27335T-A27344T-A27345T (CR5) and b) CR C10449A-T10459C (CR8), which are two CRs detected in Houston wastewater that were supported by clinical samples from Houston as well as from other US cities. For each of the CRs, the top figure shows the prevalence rate in Houston clinical samples in bars and the intra-host allele frequency of the CR in supported clinical samples in the box plot. The center heatmap shows the prevalence rate in other US regions, where white cells indicate no data. The bottom figure shows the distribution of the PANGO lineages of the consensus genomes of the CR in supported clinical samples in Houston and other US regions. All PANGO lineages of Omicron other than BA.1.1, BA.1.15, BA.1.17, BA.1.18 and BA.1.20 are combined and denoted as Omicron.

### Supplementary Tables

| | Texas | Non-Texas | Combined |
|---|---|---|---|
| **Total Sample Count** | 2,458 | 4,655 | 7,113 |
| **Supported Sample Count** | 28 | 17 | 45 |
| **Prevalence Rate** | 0.0114 | 0.0037 | 0.0063 |
| **Allele Freq Max** | 0.0417 | 0.0185 | 0.0417 |
| **Allele Freq Mean** | 0.0064 | 0.0047 | 0.0057 |
| **Collection Date Start** | 2021-11-06 | 2022-01-24 | 2021-11-06 |
| **Collection Date End** | 2022-03-21 | 2022-02-13 | 2022-03-21 |

**Supplementary Table 1. Pacbio Clinical Samples Supporting CR12 (A29039T-G29049A).** A total of 7113 Pacbio HiFi clinical samples were selected from NCBI SRA database and the sequencing reads were aligned to the reference genome of SARS-CoV-2. The table shows the distribution of samples collected in Texas and other US states containing reads supporting CR A29039T-G29049A, as well as the prevalence rate, max and mean allele frequency of the CR, and the range of sample collection dates.

| | Nucleotide Mutation | Prevalence in Wastewater Samples | Prevalence in Clinical Samples | Consensus Mutation | Translation | Sequences with the AA change | Reported Countries |
|---|---|---|---|---|---|---|---|
| CR1 | A26530G(*) | High | High | Yes | M:D3G | 2.29M | 201 |
| | C26577G(*) | High | High | Yes | M:Q19E | 7.65M | 206 |
| | G26634A | Low | Low | No | M:A38T | 15 | 8 |
| CR2 | C6402T | High | Low | Yes | ORF1a:P2046L | 3.96M | 200 |
| | G6456A | Medium | Medium | No | ORF1a:C2064Y | 436 | 29 |
| CR3 | A26530G(*) | High | High | Yes | M:D3G | 2.29M | 201 |
| | C26533T | Low | Low | No | M:S4F | 6004 | 104 |
| | C26542A | Medium | Medium | No | M:T7N | 212 | 31 |
| | T26545G(*) | Medium | Medium | No | M:I8S | 67 | 22 |
| CR4 | A26530G(*) | High | High | Yes | M:D3G | 2.29M | 201 |
| | T26545G(*) | Medium | Medium | No | M:I8S | 67 | 22 |
| CR5 | A27259C | High | High | Yes | ORF6:R20R | 5.74M | 206 |
| | C27335T | Medium | High | No | ORF6:T45I | 1632 | 67 |
| | A27344T(*) | Medium | High | No | ORF6:K48I | 154 | 26 |
| | A27345T(*) | Medium | High | No | | | |

| | | | | | | | |
|---|---|---|---|---|---|---|---|
| CR6 | T29029C | Medium | Low | No | N:A252A | 3045 | 75 |
| | A29039T | Medium | High | No | N:K256* | 118 | 12 |
| CR7 | A26530G(*) | High | High | Yes | M:D3G | 2.29M | 201 |
| | C26577G(*) | High | High | Yes | M:Q19E | 7.65M | 206 |
| | C26625A | Medium | Low | No | M:L35I | 25 | 7 |
| CR8 | C10449A | High | High | Yes | ORF1a:P3395H | 7.96M | 207 |
| | T10459C | Medium | Low | No | ORF1a:T3398T | 493 | 46 |
| CR9 | T15682A | Medium | Low | No | ORF1b:Y739N | 20 | 8 |
| | T15685A | Medium | Low | No | ORF1b:L740M | 1145 | 31 |
| CR10 | A24966T | Medium | Low | No | S:N1135I | 42 | 7 |
| | C25000T | High | High | Yes | S:D1146D | 7.90M | 207 |
| CR11 | A27344T(*) | Medium | High | No | ORF6:K48I | 154 | 26 |
| | A27345T(*) | Medium | High | No | | | |
| | A27354G | Medium | High | No | ORF6:Q51Q | 3288 | 72 |
| CR12 | A29039T(*) | Low | Medium | No | N:K256* | 118 | 12 |
| | G29049A | Low | Medium | No | N:R259Q | 153 | 34 |

**Supplementary Table 2. Individual mutations of CR1-CR12.** This table contains the information of the prevalence rate of each individual mutation of CR1-CR12 in Houston wastewater and clinical samples, as well as the corresponding amino acid changes of the mutations. We further queried each amino acid change with outbreak.info, and the table shows the number of sequences containing each of the amino acid change around the world (as of 2023-11-02), as well as the number of countries that the amino acid change has been reported. Nucleotide mutation that have been repeatedly found in multiple CRs are marked with (*).

### Supplementary Information

#### ST1. CRs may contain signal of recombination events

Most of the CRs we found in Houston wastewater contain signature mutations that are specific to only one PANGO lineage, with a few exceptions. A combination of A29301G (N:D343G) and G29402T (N: D377Y) was detected in 22 of the wastewater treatment plants between 2021-12-23 and 2022-01-06. Based on the definition of signature mutations, A29301G (N:D343G) is specific to Omicron BA.1.15, with more than 97% of the reported BA.1.15 (and its descendants BA.1.15.1, BA.1.15.2, and BA.1.15.3) genomes in GISAID EpiCoV containing such mutation. On the other hand, multiple Delta strains in the database, including AY.29.1, AY.9.21, etc, present the G29402T (N: D377Y) mutation. This rare observation may be hinting at a potential recombination event between early Omicron strains and Delta strains of SARS-CoV-2. However, confirming the emergence of recombinant strains requires an examination on mutations across much larger regions of the SARS-CoV-2 genomes, which can not be acquired via wastewater surveillance[56,57].
